# Identifying patient-level risk factors associated with non-*β*-lactam resistance outcomes in invasive methicillin-resistant *Staphylococcus aureus* infections in the United States using chain graphs

**DOI:** 10.1101/2021.11.12.21266278

**Authors:** William J. Love, Christine A. Wang, Cristina Lanzas

**Affiliations:** Department of Population Health and Pathobiology North Carolina State University, Raleigh, NC, USA

## Abstract

Methicillin-resistant *Staphylococcus aureus* (MRSA) is one of the most common causes of hospital- and community-acquired infections. MRSA is resistant to many antibiotics, including ß-lactam antibiotics, fluoroquinolones, lincosamides, macrolides, aminoglycosides, tetracyclines, and chloramphenicol. Graphical models such as chain graphs can be used to quantify and visualize the interactions among multiple resistant outcomes and their explanatory variables. In this study, we analyzed MRSA surveillance data collected by the Centers for Disease Control and Prevention (CDC) as part of the Emerging Infections Program (EIP) using chain graphs with the objective of identifying risk factors for individual phenotypic resistance outcomes (reported as minimum inhibitory concentration, MIC) while considering the correlations among themselves. Some phenotypic resistances have low connectivity to other outcomes or predictors (e.g. tetracycline, vancomycin, doxycycline, and rifampin). Levofloxacin was the only resistant associated with healthcare use. Blood culture was the most common predictor of MIC. Patients with positive blood culture had significantly increased MIC to chloramphenicol, erythromycin, gentamicin, lincomycin, and mupirocin, and decreased daptomycin and rifampin MICs. Chain graphs show the unique and common risk factors associated with resistance outcomes.

## INTRODUCTION

Antimicrobial resistance (AMR) poses a significant threat to modern medicine by decreasing the efficacy of antimicrobial treatment and increasing patient adverse outcomes and healthcare costs (Cantón & Ruiz-Garbajosa 2011, WHO 2014). Many of the bacterial pathogens of most clinical concern are resistant to multiple classes of antimicrobials and referred to as multidrug-resistant (MDR). Among MDR pathogens, methicillin-resistant *Staphylococcus aureus* (MRSA) is one of the most common causes of hospital- and community-acquired infections. In 2019, the CDC classified MRSA as a serious threat (CDC, 2019). MRSA has become resistant to many antibiotics used for its treatment besides methicillin and most other ß-lactam antibiotics, and is frequently resistant to fluoroquinolones, lincosamides, macrolides, aminoglycosides, tetracyclines, and chloramphenicol (Vestegard et al., 2019). Multidrug-resistant phenotypes often result from accumulating multiple genes encoding for resistance to different single drugs on mobile genetic elements (e.g. plasmids and integrons) and individual genes encoding multidrug efflux pumps (Nikaido, 2009; Alekshun & Levy, 2007). These genetic elements cause MDR phenotypes to be more common than would be expected if resistances were independent, and the lack of independence in resistance phenotypes allows for co-selection which can drive further changes in the prevalence of resistance within a pathogen population (Lehtinen et al., 2019). In other words, selection for one resistance may select for resistance to other unrelated antibiotic drug classes. For instance, increased prescription of tetracyclines and gentamicin in Europe during the 1980s was associated with an increased prevalence of MRSA (Hawkey, 2008).

Clinical infection with MDR pathogens results in reduced therapeutic efficacy of antibiotics and worse patient outcomes (Tacconelli, 2009). Accounting for the MDR nature of pathogens is crucial to appropriately identifying risk factors for such infections, and thus improving clinical management. Antimicrobial susceptibility testing panels for *S. aureus* and other pathogens typically include 10 to 15 antimicrobials. The complex correlation structure among the antimicrobial susceptibility testing (AST) results requires multivariate analysis methods that take into account multiple outcomes simultaneously. Use of standard statistical methods, e.g., multivariable regression, treats each outcome as independent. Multivariate methods explicitly consider multiple dependent variables, but they require *a priori* assumptions of the correlation structure among outcomes, and do not allow for the simultaneous estimation of the correlations between outcomes and the effects of predictors. Alternative methods that allow for the simultaneous estimation of outcome correlations and causal effect measures are more efficient (Getoor et al., 2004). Probabilistic graphical models (PGMs) are methods for representing complex joint distributions without *a priori* designation of outcome correlation structures (Koller & Friedman, 2009).

In PGMs, nodes represent variables of interest and edges connecting nodes represent an association between those two variables (Koller & Friedman, 2009). Bayesian networks are a type of PGM that decompose he joint distribution into a directed acyclic graph (DAG) with directed edges, in which each node is conditionally independent of its non-descendants given its parents (nodes directly connected to the node of interest) (Figure 1, A). DAGs are extensively used in epidemiology to represent causal diagrams and identify statistical models most likely to yield unbiased effect estimates (Shrier and Platt, 2008). Bayesian networks have been previously applied to resistance data from clinical bacterial cultures (Cherny et al., 2020). While Bayesian networks are powerful tools for evaluating causal associations between predictor and outcome variables, associations among resistance outcomes are more appropriately represented by undirected edges. If two resistances are correlated, the increase in one may yield an increase on the other and vice versa, so assigning directionality among correlated resistant outcomes is inappropriate. Therefore, correlated resistance outcomes are better represented with PGMs with undirected edges, also called Markov networks (Figure 1, B). The structure of a Markov network of resistance outcomes can be learned by using partial correlations coefficients followed by a penalization approach to remove the weakest edges (Love et al. 2016; Love et al. 2018). For evaluating potential risk factors for multiple resistance outcomes, a third type of PGM called chain graphs shows promise. Chain graphs are a hybrid between Bayesian and Markov networks (Figure 1, C). They contain both directed and undirected edges between nodes. This enables the identification of effect estimates between risk factors and resistances (represented by directed edges) while simultaneously accounting for joint distributions of resistance outcomes (represented by undirected edges). In this study, we analyzed MRSA surveillance data collected by the CDC-EIP using chain graphs with the objective of identifying risk factors for individual resistance outcomes while considering the correlations among phenotypic resistant outcomes.

**Figure 1:**
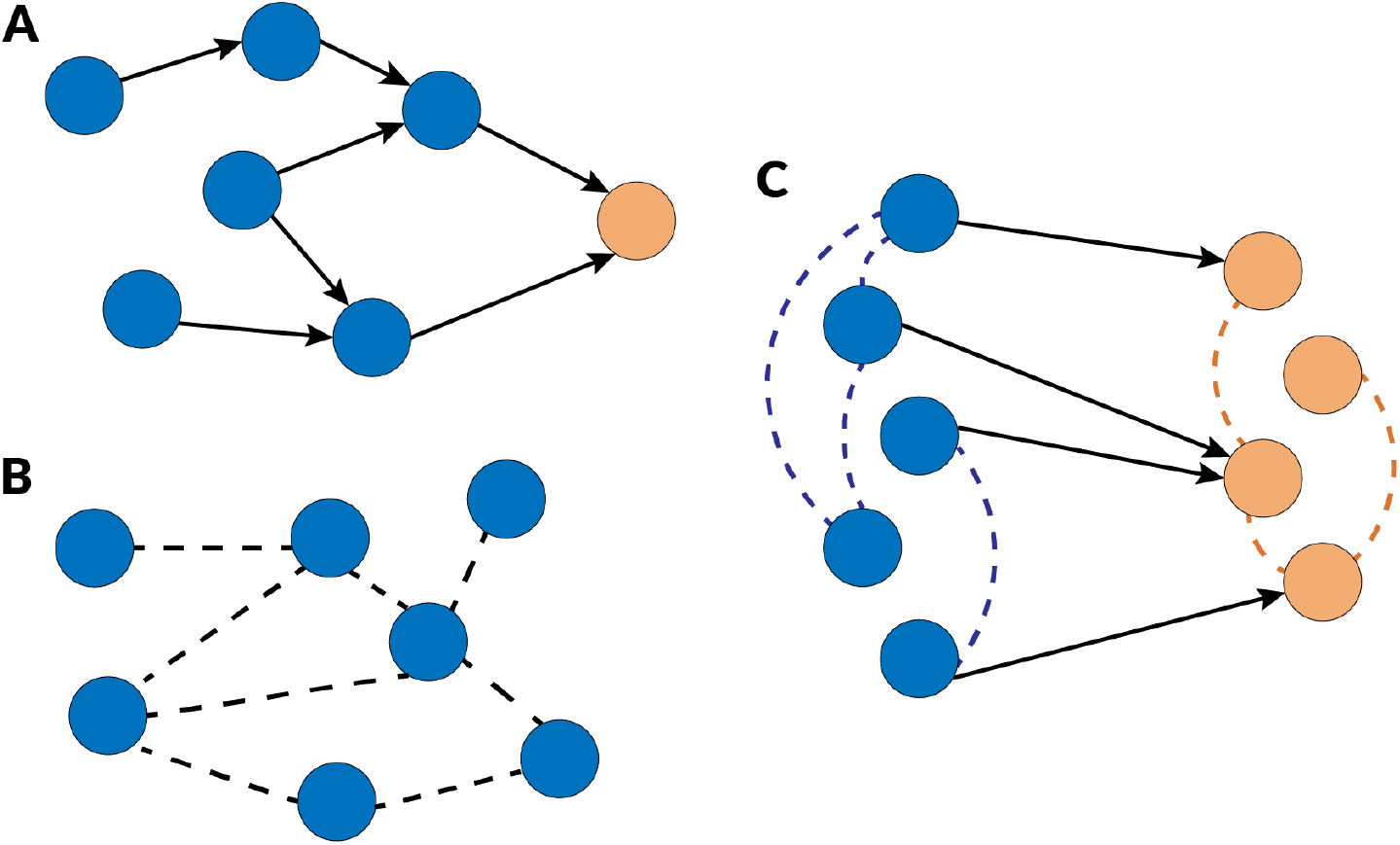
Comparison of three types of probabilistic graphical models. A) Bayesian networks have directed edges connecting nodes and are commonly used in epidemiology in the form of Directed Acyclic Graphs, which illustrate the relationship between exposure variables (blue nodes) and outcome variables (yellow node), as shown here; B) Markov networks have undirected edges connecting nodes within a graph, here depicted with a dotted line; C) chain graphs utilize both directed and undirected edges, with undirected edges connecting edges within the same layer (denoted by the respective yellow and blue color schemes) and directed edges connecting nodes between different layers.

## METHODS

### Data Description

The data set consisted of 8,982 isolates of MRSA collected from patients in 9 states between 2004 and 2016 as part of the CDC-EIP invasive *S. aureus* infection surveillance program. This program is an active population-based and laboratory-based surveillance system that monitors changes in incidence of hospital-onset (HO), healthcare-associated community-onset (HACO), and community-associated (CA) invasive infections with *S. aureus* (https://www.cdc.gov/hai/eip/saureus.html). The data included results from an AST panel of 13 antibiotics (Table 1) and 15 potential predictors describing patients’ risk factors (Table 2). HO (n = 1927) and HACO (n = 5201) isolates were both classified as healthcare-acquired. The minimum inhibitory concentration (MIC) values were log_2_ transformed so that a one unit change in the transformed value would correspond to a change of one two-fold dilution as used in microbroth dilution plates (Love et al. 2016; Hernandez and Conforti 1994). Susceptibility results which exceeded the highest tested concentration were increased by one dilution, e.g., MIC > 16 ug/mL was re-coded MIC = 32 ug/mL with log_2_(MIC) = 5.

**Table 1.**
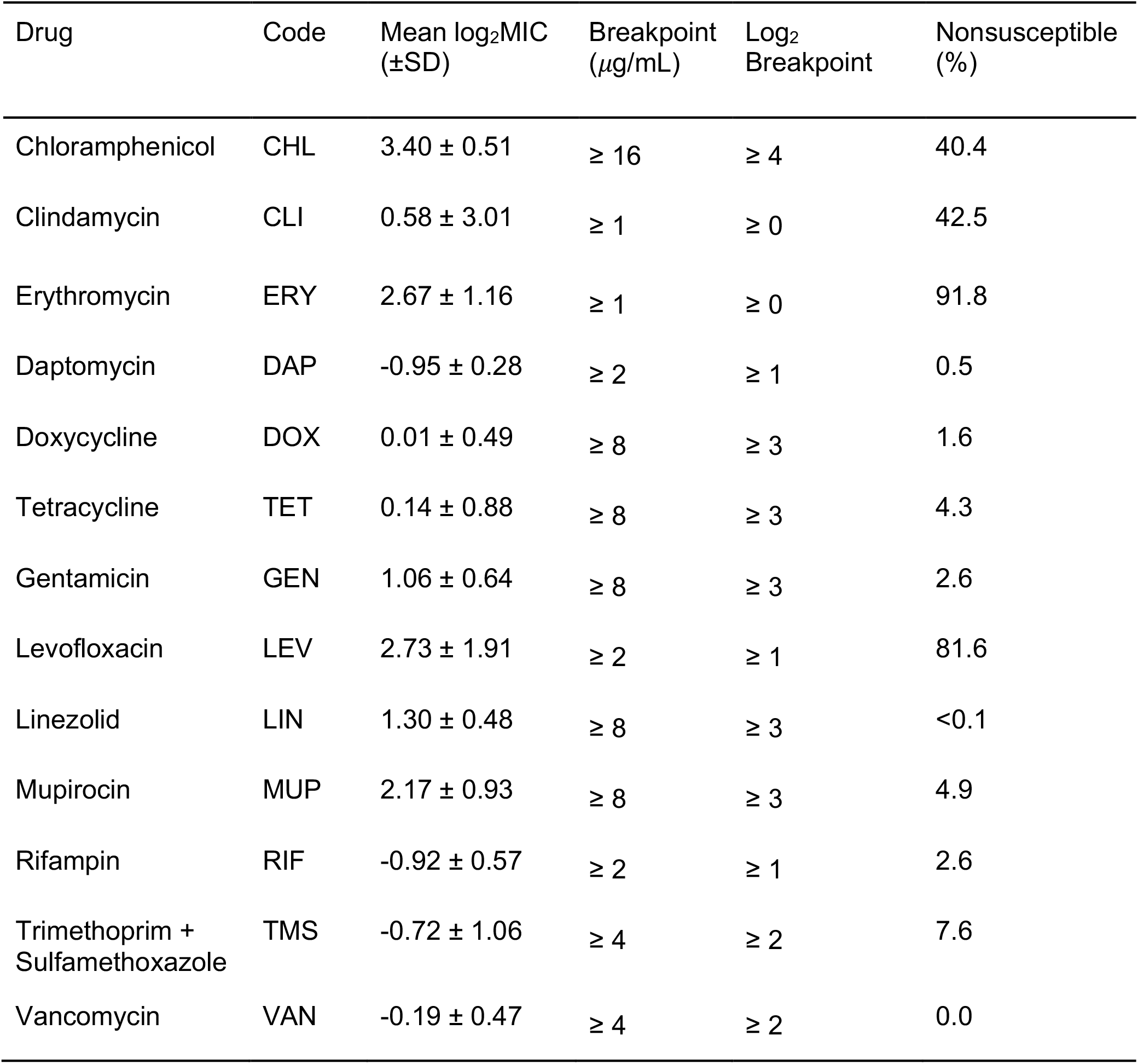
Proportions of non-susceptible isolates and breakpoints for 13 phenotypical susceptibilities in 8,982 MRSA isolates collected in the US between 2004 and 2016. Breakpoints are based on CLSI published MIC breakpoints (CLSI, 2019).

**Table 2.**
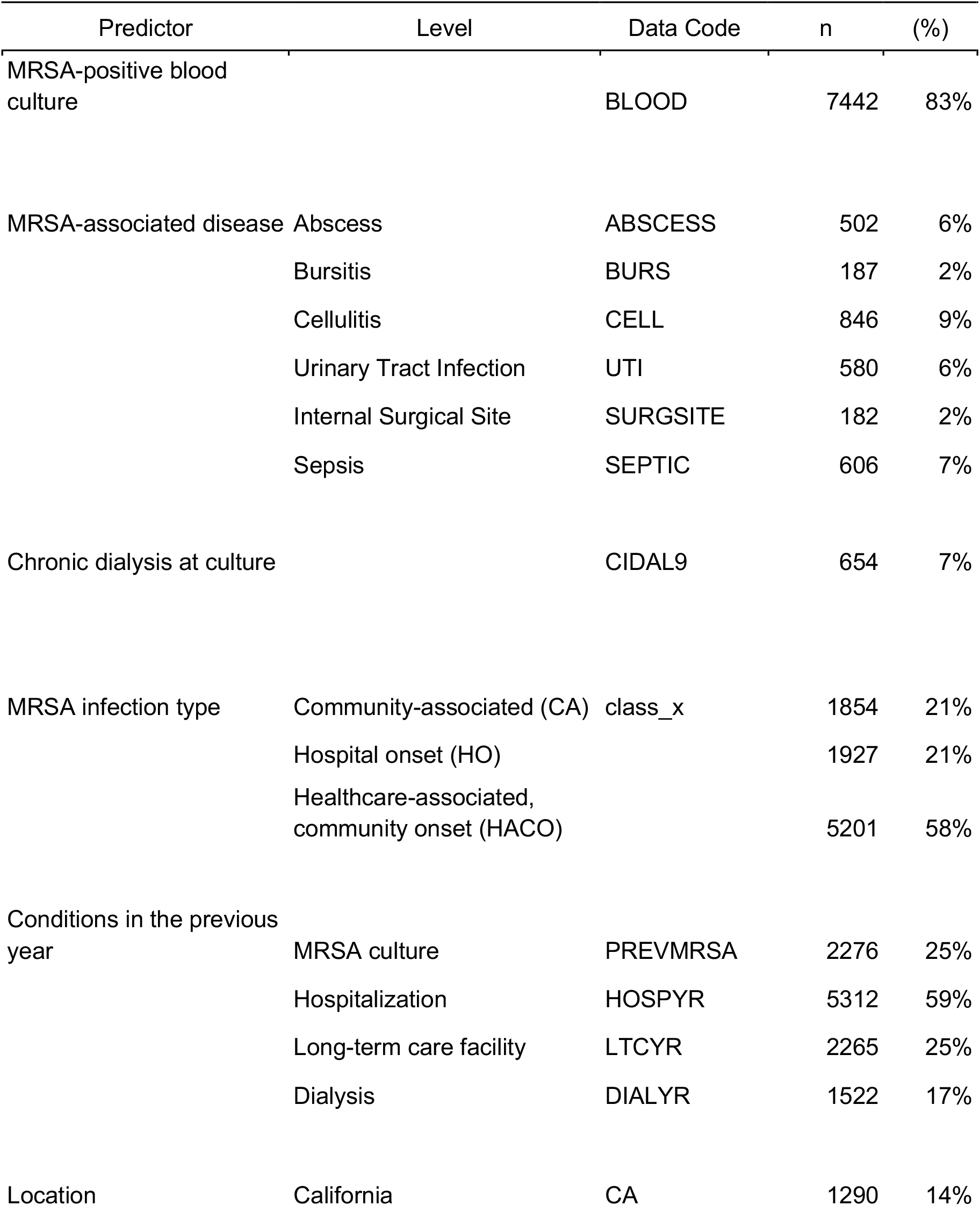
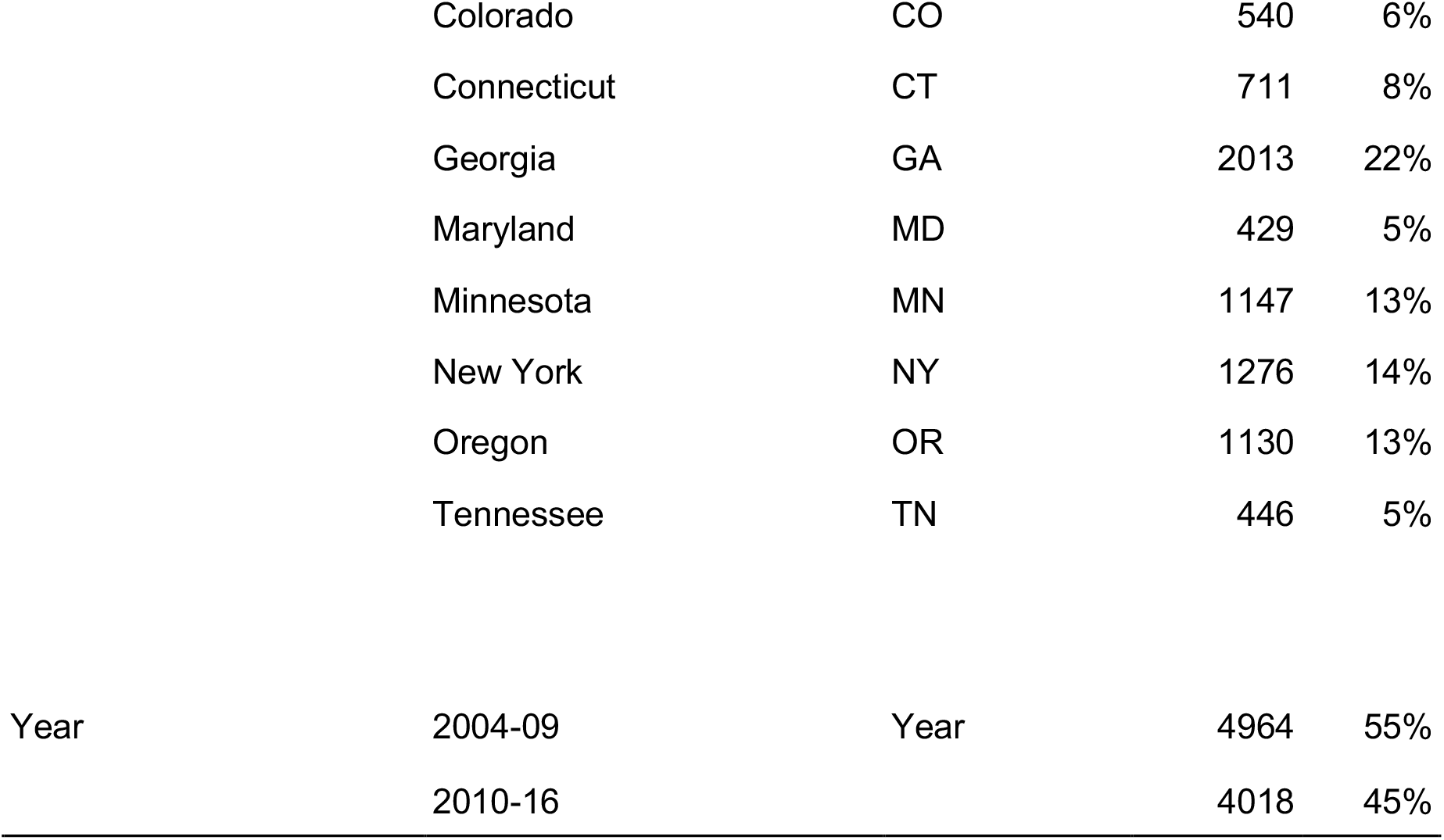
Summary of patient covariates for 8,982 MRSA isolates collected in the US between 2004 and 2016.

### Chain Graph Models

The chain graph consists of two types of nodes representing the set of outcome variables (Y), and the set of predictor variables (X) (Figure 2). The set of outcome variables are the log-transformed MICs for the 13 antibiotics of the AST panel. The undirected edges among the log_2_MIC variables represent the non-trivial partial correlations (*Ω*). The directed edges originate from the predictor variables (patient-level risk factors) towards the outcome variables (log_2_ MICs) and represent the estimated effects of predictors on the outcomes (*β*).

**Figure 2.**
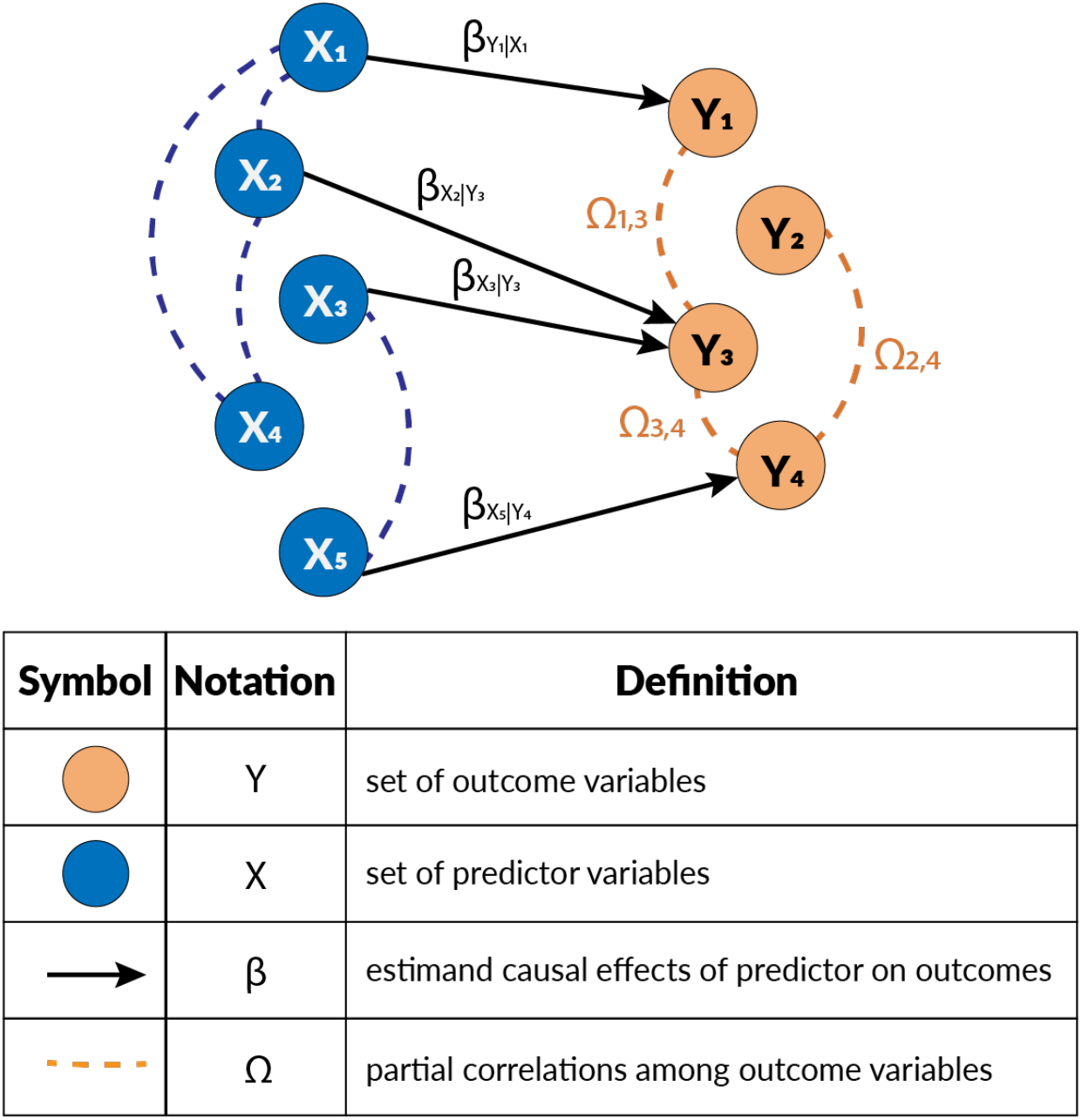
Symbology for vertices and directed and undirected components of chain graphs.

### Learning Chain Graphs from Data

Analyzing large data sets with multiple correlated outcomes presents a particular set of challenges. Very large data sets that include thousands of records can create challenges for model selection because very small effects will appear to be significant. For example, a sample size of 10,000 samples with a standard deviation of log_2_MIC equal to 3 used to make 100 comparisons will have over 95% power to identify a log_2_MIC change of 0.16 (an 11% increase in MIC) as significant with overall alpha equal to 0.05; however, a 0.16 log dilution change in average MIC is a much smaller effect than would be considered biologically important. Penalization, also called regularization, methods have been used in machine learning applications to avoid overfitting. Penalization is a soft-thresholding method that reduces trivially small effect estimates to zero (Tibshiriani 1994). Estimates that are not trivially small are biased to the null value, typically zero, *i*.*e*., the estimated magnitude of the effect is smaller than the unbiased effect. Larger penalties will reduce larger effect estimates and bias remaining estimates more towards zero and result in sparser models. Adjusting the penalty allows researchers to tune analysis in an objective manner similar to adjusting the significance level *α*. The least absolute shrinkage and selection operator (lasso) (Tibshiriani 1994) and the graphical lasso (Friedman, Hastie, and Tibshiriani 2008) are very commonly used penalization methods. To learn the graph structure and parameters, the data were fitted to chain graph models using the penalized maximum likelihood estimation method proposed by Lin et al. 2016. The directed edges that compose *β* are estimated using the debiased lasso with the penalty *λ* (Figure 2). Directed edges exist where effect estimates are not penalized to zero. The undirected edges that compose *Ω* are estimated using the graphical lasso estimated with the penalty *ρ*. The algorithm in Lin et al. 2016 has the following steps; first, all possible conditional relationships, i.e., the terms in *β*, are screened by regressing each outcome on the full set of predictors using the de-biased lasso (Javanmard and Montanari 2014). The de-biased lasso corrects for the penalization and estimates an unbiased p-value. Only terms with p < *α* are considered for inclusion in the full model. The algorithm then finds initial estimates for the *β* terms, then applies the graphical lasso to the residuals to estimate the partial correlations that comprise *Ω*. The algorithm returns to estimate *β* conditioned on the newly estimated *Ω*, and then again estimates *Ω* from the residuals. The algorithm iterates in this way, estimating *β* conditioned on *Ω* then *Ω* from the residuals until fit converges.

The stability approach to regularization selection (StARS) was used to identify a penalty pair values (*λ, ρ*) that produced the most stable set of edges (Liu, Roeder, and Wasserman 2010). The StARS method takes repeated subsamples of the data, estimates the chain graph for each subsample, and estimates the instability of edge presence across those subsamples. The instability score *S* was calculated as *S* = ∑_*i*_ 2*π*_*i*_(1 − *π*_*i*_) where *π*_i_ is the proportion of subsamples where the i^th^ edge appeared over the entire set of possible directed and undirected edges. All combinations of values between 0.10 and 0.40 in increments 0.01 were tested for *λ* and *ρ* each. The models created with penalties below 0.10 were found to be too dense to be easily interpretable, and penalties above 0.40 produced models with too few edges to be informative. Twenty subsamples of 80% of the usable data were taken for each combination. The penalty pair that minimized *S* was selected as the final model. Using the final model, the standard errors for the effect estimates were calculated using a modified ordinary least squares method to estimate residual mean square errors.

## RESULTS

The highest proportion of isolates with MICs exceeding the susceptible breakpoints were 40.4% for CHL, 42.5% for CLI, 91.8% for ERY, and 81.6% for LEV (Table 1). No other phenotypic resistances were found in more than 10% of isolates. No isolates were vancomycin-resistant and 3 isolates had intermediate susceptibility to VAN (MIC_VAN_ = 4)g/ml).

Stability selection found the most stable model structure occurred with penalties *λ* = 0.20 and *ρ*)= 0.30. Figure 3 presents a representation of the chain graph model. The model’s undirected *Ω* component had 13 edges (16.7% undirected subgraph density) and the directed *β* component had 54 edges (18.1% directed subgraph density). The degree of a node (d) describes the number of edges connected to the node. In chain graphs, the total degree (d_tot_) of the outcome variables can be split into d_B_ and d_Y_; d_B_ describes in-degree of directed edges in *β* that point at an outcome node and d_Y_ describes the number of connected undirected edges from *Ω* (Table 3). To facilitate the visualization of the chain graph, Figure 4 breaks the overall chain graph into the neighbourhood graphs. Each outcome node is displayed with its adjacent nodes. Overall, the total degree was highly variable across resistance outcomes. VAN and TET had only one edge, while CHL had a degree of 14 (Table 3).

**Table 3.** Degree (d) of outcome nodes (Y) in the selected chain graph model, divided into in-degree from predictors on outcomes (d_B_), undirected degree for other outcome nodes (d_Y_),and total degree (d_B_ + d_Y_ = d_tot_).

| Y | $d_B$ | $d_Y$ | $d_{tot}$ |
| --- | --- | --- | --- |
| CHL | 8 | 6 | 14 |
| CLI | 5 | 1 | 6 |
| DAP | 1 | 4 | 5 |
| DOX | 0 | 2 | 2 |
| ERY | 9 | 1 | 10 |
| GEN | 3 | 3 | 6 |
| LEV | 8 | 1 | 9 |
| LIN | 3 | 2 | 5 |
| MUP | 6 | 3 | 9 |
| RIF | 2 | 2 | 4 |
| TET | 0 | 1 | 1 |
| TMS | 8 | 0 | 8 |
| VAN | 1 | 0 | 1 |

**Figure 3.**
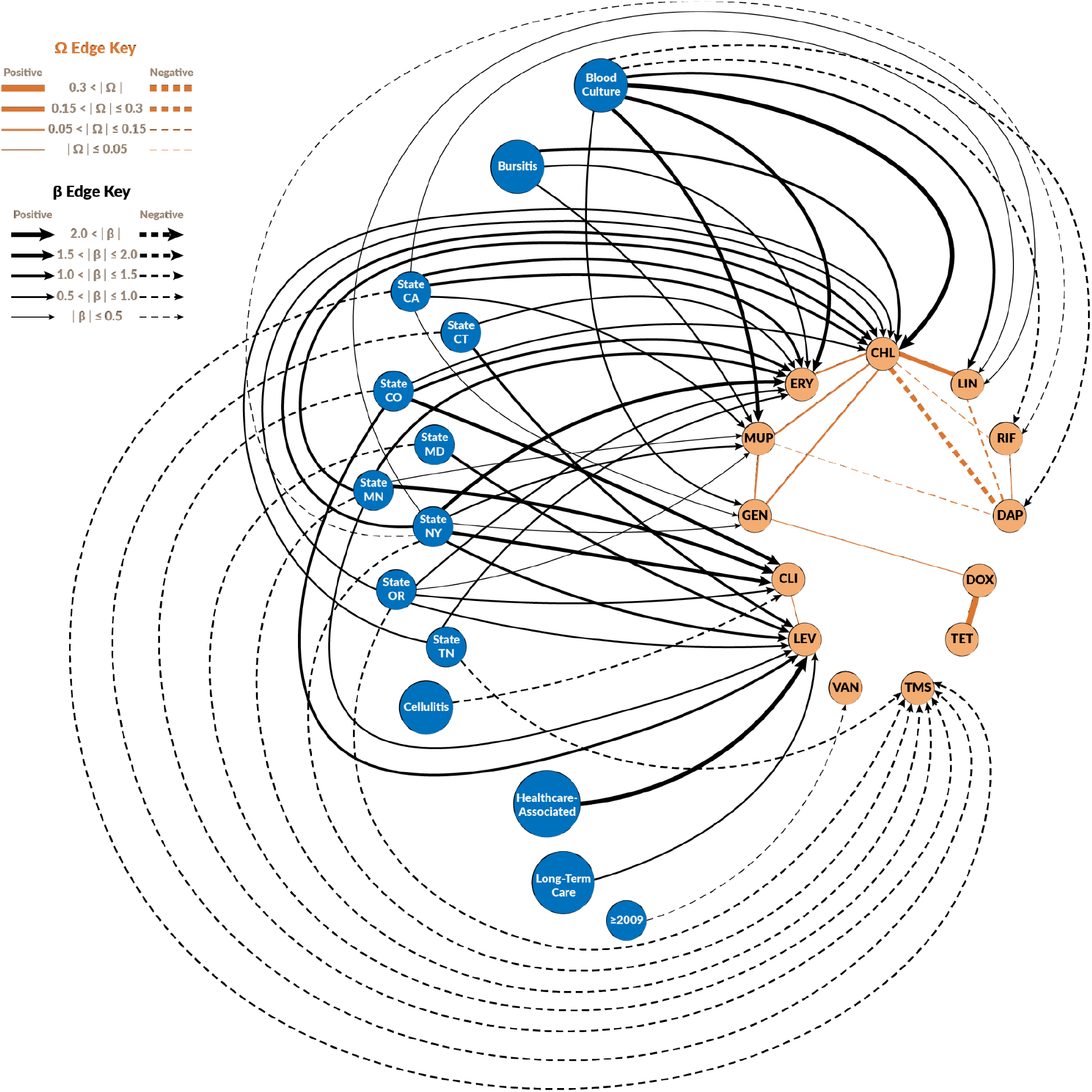
Chain graph representation of the CDC-EIP invasive methicillin-resistant *Staphylococcus aureus* surveillance data. Orange nodes represent the resistance outcomes (as log_2_MIC) and the blue nodes are the risk factors associated with the log_2_MIC nodes. The edges are decorated with line weights and styles to indicate the magnitude of the partial correlations among resistances (*Ω*) and the magnitude of the effect estimates (*β*).

**Figure 4.**
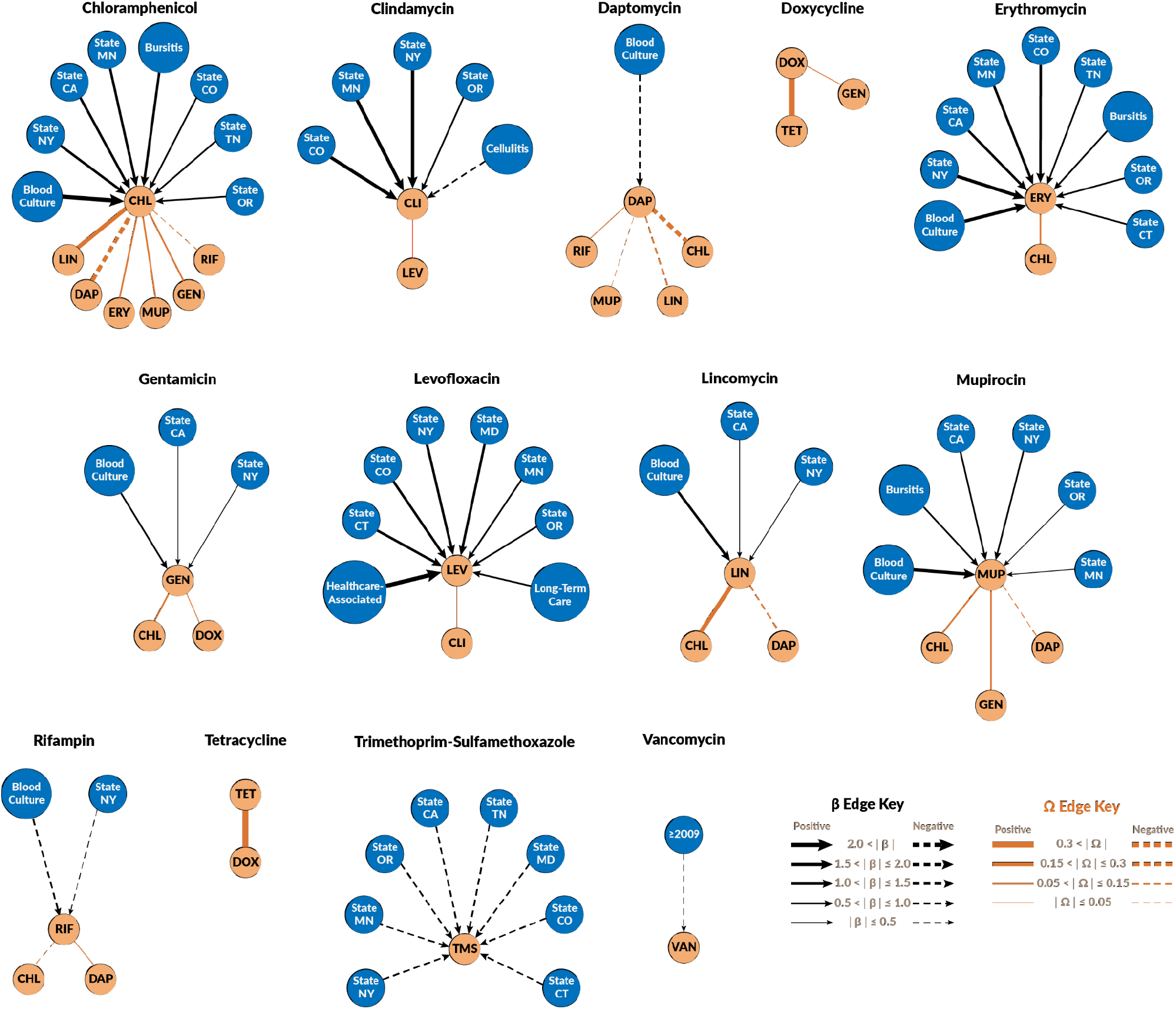
Neighbourhood graphs for the chain graph representing the CDC-EIP invasive methicillin-resistant *Staphylococcus aureus* surveillance data. Orange nodes represent the resistance outcomes (as log_2_MIC) and the blue nodes are the risk factors associated with the log_2_MIC nodes. The edges are decorated with line weights and and styles to indicate the magnitude of the partial correlations among resistances (*Ω*) and the magnitude of the effect estimates (*β*).

The directed edges in the graphical model represent the de-biased non-zero estimated linear coefficients describing the average effect measure of the patient predictors on the outcome MICs and are summarized in Table 4. Eight of the fifteen predictors summarized in Table 2 had no significant relationship with any of the assayed susceptibilities. Eleven of thirteen susceptibilities were affected by at least one predictor and nine of the susceptibilities had some degree of regional variance (Table 4). The undirected edges in the graphical model represent the penalized partial correlations between the outcome MICs and are summarized in Table 5.

**Table 4.** Directed effect estimates (*β*) of patient factors (X) on log_2_ susceptibilities (Y) from the chain graph model built 8,962 MRSA isolates.

| Y | X | $\beta$ | Std. Err. |
| --- | --- | --- | --- |
| CHL | MRSA-positive blood culture | 2.72 | 0.03 |
| CHL | MRSA-associated bursitis | 1.02 | 0.08 |
| CHL | California | 1.22 | 0.04 |
| CHL | Colorado | 0.95 | 0.05 |
| CHL | Minnesota | 1.05 | 0.04 |
| CHL | New York | 1.42 | 0.04 |
| CHL | Oregon | 0.68 | 0.04 |
| CHL | Tennessee | 0.71 | 0.05 |
| CLI | MRSA-associated cellulitis | -0.73 | 0.10 |
| CLI | Colorado | 1.88 | 0.13 |
| CLI | Minnesota | 1.65 | 0.09 |
| CLI | New York | 1.57 | 0.09 |
| CLI | Oregon | 0.75 | 0.09 |
| ERY | MRSA-positive blood culture | 1.86 | 0.04 |
| ERY | MRSA-associated bursitis | 0.95 | 0.10 |
| ERY | California | 1.34 | 0.05 |
| ERY | Colorado | 1.12 | 0.06 |
| ERY | Connecticut | 0.82 | 0.06 |
| ERY | Minnesota | 1.12 | 0.05 |
| ERY | New York | 1.51 | 0.05 |
| ERY | Oregon | 0.94 | 0.05 |
| ERY | Tennessee | 0.96 | 0.07 |
| DAP | MRSA-positive blood culture | -0.90 | 0.01 |
| GEN | MRSA-positive blood culture | 0.97 | 0.02 |
| GEN | California | 0.40 | 0.02 |
| GEN | New York | 0.33 | 0.02 |
| LEV | Long-term care residence in the previous year | 0.56 | 0.05 |
| LEV | HO/HACO MRSA | 2.20 | 0.05 |
| LEV | Colorado | 1.33 | 0.08 |
| LEV | Connecticut | 1.35 | 0.07 |
| LEV | Maryland | 1.15 | 0.09 |
| LEV | Minnesota | 0.97 | 0.06 |
| LEV | New York | 1.21 | 0.06 |
| LEV | Oregon | 0.73 | 0.06 |
| LIN | MRSA-positive blood culture | 1.19 | 0.02 |
| LIN | California | 0.28 | 0.02 |
| LIN | New York | 0.41 | 0.02 |
| MUP | MRSA-positive blood culture | 1.88 | 0.03 |
| MUP | MRSA-associated bursitis | 0.89 | 0.09 |
| MUP | California | 0.74 | 0.04 |
| MUP | Minnesota | 0.42 | 0.04 |
| MUP | New York | 0.69 | 0.04 |
| MUP | Oregon | 0.48 | 0.04 |
| RIF | MRSA-positive blood culture | -0.87 | 0.02 |
| RIF | New York | -0.33 | 0.02 |
| TMS | California | -0.91 | 0.04 |
| TMS | Colorado | -0.84 | 0.05 |
| TMS | Connecticut | -0.82 | 0.04 |
| TMS | Maryland | -0.89 | 0.05 |
| TMS | Minnesota | -0.96 | 0.04 |
| TMS | New York | -0.99 | 0.04 |
| TMS | Oregon | -0.95 | 0.04 |
| TMS | Tennessee | -0.90 | 0.05 |
| VAN | ≥ 2009 | -0.42 | 0.01 |

**Table 5.** Penalized partial correlation estimates (*ω*)) from the chain graph model built 8,962 MRSA isolates.

| Susceptibilities | | $\omega$ |
| --- | --- | --- |
| CHL | ERY | 0.14 |
| CHL | DAP | -0.19 |
| CHL | GEN | 0.06 |
| CHL | LIN | 0.26 |
| CHL | MUP | 0.14 |
| CHL | RIF | -0.04 |
| CLI | LEV | 0.05 |
| DAP | LIN | -0.07 |
| DAP | MUP | -0.02 |
| DAP | RIF | < 0.01 |
| DOX | TET | 0.43 |
| DOX | GEN | 0.01 |
| GEN | MUP | 0.11 |

The model found two susceptibilities (TET and DOX) without any significantly associated covariates (Figure 4, Table 4). Patients with positive blood cultures had significantly increased MICs (*α*=0.05) for five tested susceptibilities (*β*_CHL|BLOOD_ = 2.72, *β*_ERY|BLOOD_ = 1.86, *β*_GEN|BLOOD_ = 0.97, *β*_LIN|BLOOD_ = 1.19, *β*_MUP|BLOOD_ = 1.88) and significantly decreased in two (*β*_DAP|BLOOD_ = −0.90, *β*_RIF|BLOOD_ = −0.87). On average, samples from 2009 and later had significantly lower VAN (*β*_VAN|yr>2009_ = −0.42). Significant variation by state of origin was found in all tested susceptibilities except VAN and DAP. Patients with MRSA-associated abscesses, UTIs, infected internal surgical sites, sepsis, previous diagnosis of MRSA, dialysis the previous year, or current chronic dialysis were not significantly associated with any susceptibilities. Fitted values for each observation were back calculated and used to estimate mean square error and approximate p-value for each parameter. The least significant Z-score was approximately −7.0 with p = 2.7 × 10^−12^. Of the 54 estimated parameters, all but two had p-values less than 2.2 × 10^−16^, the smallest double value R’s precision can represent.

## DISCUSSION

The chain graph model fit to the surveillance data showed complex and varied relationships between patient predictors and measured susceptibilities. Some of the resistances on the panel, such as TMS and VAN, were each only affected by a single predictor (region and time, respectively), and were independent of all other resistances (Figure 4). Other resistances, such as CHL, were affected by multiple predictors and were correlated with many other resistances (Figure 4). It appears likely that within the sampled population of MRSA isolates, different types of drivers are responsible for the resistance phenotypes noted.

IDSA guidelines recommend clindamycin, combination sulfonamides, and doxycycline to treat uncomplicated skin and soft tissue MRSA infections (Liu et al., 2011). DOX was associated with TET ()_DOX-TET_ = 0.43) and weakly with GEN ()_DOX-GEN_ = 0.01). DOX and TET had the highest partial correlation among resistances, which would be expected given the structural and functional similarity between both drugs. Gentamicin and doxycycline both bind to the 30S ribosomal subunit, so some mutations may affect both resistance simultaneously. TMS showed only regional variation, with all eight states other than GA having average TMS just under one dilution less than the isolates from GA. Isolates from patients with MRSA-associated cellulitis had slightly lower CLI on average (*β*_CLI|Cellulitis_ = −0.76) and showed some regional variation with four states. The only resistance associated with CLI was LEV ()_CLI-LEV_ = 0.05), so fluoroquinolone use may exert a weak indirect selection pressure for increased CLI.

Vancomycin is commonly recommended as treatment for a variety of MRSA infections including bacteremia, endocarditis, pneumonia or bone and joint infections (Liu et al., 2011). VAN was largely independent of other factors in the study. Only year was a meaningful predictor of VAN with evidence that resistance has gone down slightly in isolates collected after 2009 compared to those collected in 2008 or earlier (*β*_VAN|year_ = −0.42), and nearly all of this variation was within the susceptible range of MICs. Other resistances were not associated with VAN. Despite prior reports that *in vitro* exposure to vancomycin may select for higher daptomycin MIC (Patel et al., 2006), there was no association between both MICs in our analysis. Resistance to daptomycin was found to be common among MRSA isolates with vancomycin resistance, but the EIP data had no isolates with resistance or intermediate MIC values for vancomycin to begin with (Table 1).

Other drugs used to treat MRSA cases include linezolid, rifampicin, and daptomycin (Liu et al., 2011). Overall, the penalized partial correlations between these MICs and other resistances were small, except for correlations with CHL which are discussed below. Very weak negative partial correlations were found between DAP and LIN ()_DAP-LIN_ = −0.07) and MUP ()_DAP-MUP_ = −0.02). A very weak positive partial correlation with RIF()_DAP-RIF_ < 0.01) was noted, meaning that rifampicin and daptomycin use are unlikely to meaningfully impact resistances to each other. DAP was lower in patients with positive MRSA blood cultures (*β*_DAP|BLOOD_ = −0.90).

Chloramphenicol MIC appeared to be the most connected resistance in the estimated graphical model with d_tot_ = 14. CHL was adjacent to 6 other resistances: ERY, DAP, GEN, LIN, MUP, RIF. Chloramphenicol exposure is not a significant selective pressure on this MRSA population, which may explain the existent variance (and edges) between CHL and other resistances. CHL may be weakly selected for or against by other resistances, but not from chloramphenicol exposure. While 40.4% of the isolates had CHL in excess of the susceptible breakpoint, most of these isolates MICs fell into CLSI’s intermediate range (16)g/ml) for chloramphenicol and only 37 (0.4%) isolates had MICs that met or exceeded of the resistant breakpoint ()32)g/ml) (CLSI, 2019). Only one isolate was resistant to chloramphenicol and linezolid. The low MICs for CHL and LIN imply genes which confer strong resistance to CHL and LIN, e.g., chloramphenicol-specific acetyltransferases (*catA, catB*) (Schwarz, et al, 2004), rRNA methylation (*cfr*) that confers resistance to multiple antimicrobial classes (Shen et, al, 2013; Long et al 2015), some ATP-binding cassette (ABC) transporters (*OptrA*) that confers resistance to phenicols and oxazolidinone (Sharkey, et al, 2016; Wilson, 2016; Wang, et al, 2018), or phenicol-specific efflux pumps (*fexA, fexB*) (Kehrenberg and Schwarz, 2004) likely do not play an important role in these correlations.

The negative correlations between CHL and RIF and DAP are likely attributable to difference in strains, where determinants of chloramphenicol susceptibility do not coincide with determinants of rifampicin susceptibility, e.g., *rpoB* (Goldstein, 2014) or determinants of daptomycin susceptibility, e.g., *dltABCD, mprF*, and others (Tran, et al, 2016). Susceptibility to mupirocin can be conferred via the genese *mupA* and *mupB* and via mutations to its target isoleucyl-tRNA encoded by *ileS*. These mechanisms do not appear to provide cross-resistance to other antimicrobial classes, but *MupA* and *mupB* can reside on plasmids known to carry other susceptibility determinants (Rahman, et al 1989, Morton, et al, 1995). However, no references describing plasmids carrying both chloramphenicol and mupirocin resistance genes could be found. One possible explanation for the remainder of the CHL neighborhood could be multi-resistance plasmids similar to pSCFS1 (Kehenberg, et al 2004). This plasmid originally found in *S. sciuri* harbors a *lsa*(B) which is a different ATP-binding cassette transporter that induces low-level linezolid resistance, *erm*(33) which is a rRNA methylase that induces resistance to MLS antibiotics. In this case the plasmid could harbor another gene that induces low-to-moderate chloramphenicol resistance such as a less effective *cat* or *fex* gene, instead of the *cfr* gene identified on pSCFS1. This combination of genes on a resistance plasmid would explain the moderate to strong positive partial correlations observed between CHL and LIN (0.26) and ERY (0.14), all of which target the 50s ribosomal subunit. The difficulty in identifying the mechanisms responsible for the observed MIC correlations with CHL in this population of MRSA demonstrates the limitations of using phenotypic data alone to interpret noted patterns of resistance in complex cases. Further work is needed to incorporate genetic and genomic information into the chain graphs to better understand the molecular mechanisms responsible for the network patterns observed.

In this surveillance data, 36.8% of the CA-MRSA isolates and 13.9% of HA-MRSA isolates were susceptible to levofloxacin. Healthcare-associated MRSA isolates had average LEV values more than two dilutions higher than community associated infections (*β*_LEV|hosp_ = 2.20), and patients with a history of hospitalization in the previous year had a 0.56 dilution increase in LEV on average. The low prevalence of levofloxacin susceptibility is consistent with previous studies of multi-drug resistant MRSA in which fluoroquinolones resistance was common in HA-MRSA clones (Knight et al. 2012) and resistance rapidly evolved when fluoroquinolones were used to treat MRSA (Blumberg et al. 1991). We suspect that patients with a history of hospitalization or healthcare-associated infections were more likely to be prescribed fluoroquinolones to treat their MRSA-associated conditions or other infections or to have come into contact with MRSA isolates that have experienced selection for reduced fluoroquinolone susceptibility. However, this cannot be tested without patients’ prescription histories or information about the isolates where patients were hospitalized. Previous studies have identified fluoroquinolone treatment as a risk factor for MRSA colonization, but not MSSA colonization (Crowcroft et al. 1999; Weber et al. 2003). This is consistent with expected selection of MRSA by fluoroquinolone prescription, but it cannot be tested without information about contemporary MSSA isolates or comparable patients not colonized with *S. aureus*.

The MRSA clonal groups found in healthcare-associated infections are generally thought to be distinct from those responsible for community-acquired infections (Lakhundi and Zhang, 2018). However, LEV was the only phenotypic susceptibility found to be different between healthcare- and community-associated isolates. HA-MRSA strains tend to be more multidrug resistant because the integrons found in HA-MRSA often contain several gene cassettes and resistance genes (Lakhundi and Zhang, 2018). Three plausible explanations for why HA-vs CA-origin was not identified as a risk factor in our analysis with the exception of LEV are 1) that the epidemiological definition of HA-MRSA and CA-MRSA resulted in misclassified strains, 2) that the CA-MRSA strains are also likely to contain additional resistances acquired through plasmids or 3) that similar strains are circulating in both health-care settings and the community (Lakhundi and Zhang, 2018). USA300, a strain originally linked to CA-MRSA, is also identified as source of HA-MRSA (See et al., 2020).

Patients with positive blood cultures had significantly higher average CHL, ERY, GEN, LIN and MUP values and lower average DAP and RIF. Bacteremia is a significant source of morbidity and mortality (CDC 2019; WHO 2014) in MRSA infections. Standard of care for treating MRSA bacteremia is daptomycin and vancomycin (Liu et al., 2011). In this study, DAP and RIF were actually lower in bacteremic patients and MICs for these antimicrobials were not significantly different in bacteremic patients. These findings suggest that the recommended practices remain sound for promoting efficacy of rifampin and daptomycin.

There are several limitations in this study. The partial correlations among phenotypic resistance that remain after accounting for the patient-level risk factors can be due to multiple factors. Prescription patterns, community patterns of comorbid illness, genetic structure of MRSA isolates can all influence how drug resistances interplay (Andreatos et al., 2018, Jamrozy et al., 2016). Without additional information (e.g. sequences), these factors cannot be untangled given the available data. In addition, EIP surveillance reports selected MRSA cases from a few states, and therefore it is not necessarily generalizable to the U.S. population.

There have been a few previous studies that have approached AMR as a multivariate problem, but they have not applied a chain graph or comparable approach. One previous study applied principal component analysis and factor analysis to identify groups of antibiotics with similar trends in MIC from 15 groups of microbes from 17 studies (Hernandez and Conforti 1994). Rotated principal components correspond to dense subregions in probabilistic graphical models, *i*.*e*., groups of variables that load heavily on rotated principal components will also tend to have more edges between them in PGMs than would be expected by the overall graphical density (Love et al. 2018). The PGMs give a much more detailed picture of how the variables are correlated and do not enforce or require assumptions of orthogonality. Bayesian networks have been previously applied to identify risk factors for multiple resistant outcomes (Cherny et al., 2021, Hartnack et al., 2019). Cherny et al. (2021) investigated patient demographic and prior antibiotic use as risk factors for resistant outcomes for five different healthcare-associated pathogens. Several strong positive associations among resistances were identified for all five pathogens (Cherny et al., 2011). Chain graphs are a new analytical approach that can provide insights on the interplay between multiple resistance outcomes and their risk factors.

## Data Availability

Data used in this study is complied and housed by the Emerging Infectious Program Healthcare-Associated Infection/Community Interface Activity (EIP HAIC), Epidemiology Research and Innovations Branch, Division of Healthcare Quality Promotion, National Center for Emerging and Zoonotic Infectious Diseases, Centers for Disease Control and Preventions, and are not publicly available. Questions regarding the availability of the data should be refer to the EIP HAIC staff.

## Acknowledgments

This work was supported by the U.S. National Institutes of Health (NIH) (R35GM134934 and F30OD30022) and the Centers for Disease Control and Prevention (CDC) (U01CK000587). The funders had no role in study design, data collection and analysis, decision to publish, or preparation of the manuscript. We thank Runa H. Gokhale, Amy Gargis, Isaac See, and Joseph Lutgring from the CDC for providing useful comments.

